# Advanced MRI scan acquisition metrics improve baseline disease severity predictions compared to traditional community MRI scan metrics

**DOI:** 10.1101/2022.07.06.22277151

**Authors:** Abdul Al-Shawwa, Kalum Ost, David Cadotte, David Anderson, Nathan Evaniew, Bradley Jacobs, Julien Cohen-Adad

## Abstract

Degenerative Cervical Myelopathy (DCM) is the functional derangement of the spinal cord and acts as one of the most common atraumatic spinal cord injuries. Magnetic resonance imaging (MRI) are key in confirming the diagnosis of DCM in patients, though the utilization of higher fidelity magnetic resonance imaging scans and their integration into machine learning models remains largely unexplored. This study looks at the predictive ability of common community MRI scans in comparison to high fidelity scans in disease diagnosis. We hypothesize that the utilization of higher fidelity “advanced” MRI scans will increase the effectiveness of machine learning models predicting DCM severity. Through the utilization of Random Forest Classifiers, we have been able to predict disease severity with 41.8% accuracy in current community MRI scans and 63.9% in the advanced MRI scans. Furthermore, across the different predictive model variations tested, the advanced MRI scans consistently produced higher prediction accuracies compared to the community MRI counterparts. These results support our hypothesis and indicate that machine learning models have the potential to predict disease severity. However, neither performed well enough to be considered for use in clinical practice, indicating that the utilization of more sophisticated machine models may be required for these purposes.

## Introduction

Degenerative Cervical Myelopathy (DCM), which includes Cervical Spondylotic Myelopathy (CSM), is described as *a functional derangement of the spinal cord*. It has been discussed in literature starting in the mid 1800s^1^, with further scientific inquiry being conducted in the mid 1900’s^2^. The widespread adoption of magnetic resonance imaging (MRI) allowed for confirmation of clinical diagnosis and the development of sophisticated surgical treatment options^3^. CSM occurs following chronic compression of the cervical spinal cord, and is recognized as the most common form of atraumatic spinal cord injury globally^4^. It typically results in symptoms of gait imbalance, pain, loss of hand function, and/or numbness^5^.

Surgery is the only effective treatment for CSM that is symptomatically progressive or at least moderate in severity, with the aim of halting the progression of symptoms and relieving spinal cord compression. This is thought to limit ongoing damage to the spinal cord, in the form of demyelination, axonal damage or ischemia.^6^ Surgery comes with risks, including both short and long-term complications. These complications are unique for each individual patient^7^, and can include dysphagia, infection, and potential neurological and/or functional decline^8^.

In this study, we aim to develop and test machine learning models tasked with predicting disease severity at the time of diagnosis through the use of MRI-derived quantitative metrics. We modelled the experience of patients who received a diagnosis of CSM and we utilized community based MRI (cMRI) sequences of the cervical spine and compared them to an advanced imaging protocol (aMRI) that was developed to evaluate chronic spinal cord injury^9–11^. The aMRI dataset included advanced imaging biomarkers of disease severity which are actively being considered for diagnostic and prognostic abilities^10^, whose effectiveness has yet to be determined.

## 2. Methods

### Study Subjects

The University of Calgary Spine Program is a group of fellowship trained surgeons including both neurosurgical or orthopedic backgrounds caring for a population of approximately 2 million persons in Southern Alberta. Since 2016, we have enrolled patients presenting to our clinics with a diagnosis of DCM into the Canadian Spine Outcomes Research Network registry, approved by our institutional ethics review board (ethics certificate number REB-15-1332). As of October 2021, our group had enrolled 394 patients into this registry and collected baseline and longitudinal characteristics previously established to be informative of CSM severity.^7^ Each patient provided written consent to participate in our national research effort and is followed by our research team for 10 years or until the patient withdraws from the study. Patients present via referral from a primary care physician in Southern Alberta with a cervical spine MRI obtained from the community. The surgeon then confirms the diagnosis and offers a management plan, which include either clinical observation or surgical intervention. Patients are offered enrollment into our registry under either the observational or surgical arm provided they do not meet any exclusion criteria. These criteria include previous cervical spinal surgery, concomitant neurological disease (such as multiple sclerosis or stroke), and a new MRI diagnosis of neoplasm or infection. We refer to this dataset as the ‘community MRI (cMRI)’ dataset. A full breakdown of the various testing protocols for the community MRI dataset used in this study can be found in our research centers previously published work.^12^

A subset of the above listed patients were enrolled in a concurrent longitudinal study titled “Personalized decision making after a diagnosis of cervical myelopathy: quantitative MRI within an artificial intelligence framework”. This study has been approved by our institutional research ethics board (certificate number REB-18-1614), and each patient provided additional consent for the collection of an advanced MRI protocol that includes diffusion tensor imaging (DTI), magnetization transfer (MT), and T_2_*-weighted image acquisitions in addition to an updated T_2_-weighted scan. We refer to this dataset as the ‘advanced MRI (aMRI) dataset’. All aMRI data was acquired on a 3T GE scanner. Subjects were carefully positioned to limit head movement and requested to not move. A T_2_-weighted acquisition was acquired as follows: FIESTA-C sequence (T_2_-weighted); 512 × 512, NEX 1.0, FOV 200 mm, slice thickness 0.4 mm; resulting in a voxel size of 0.4×0.4×0.4 mm^3^, collected in the coronal plane. The total scan time was approximately 5 minutes per patient.

aMRI records also contain Diffusion tensor imaging (DTI), magnetization transfer (MT), and T_2_*-weighted image acquisitions, each with 13 axial slices across the C1 to C7 vertebrae, using a variable gap to alternate between bone and intervertebral discs. DTI used spin-echo single shot echo planar imaging (ssEPI) with 3 acquisitions averaged offline, b = 800 s/mm^2^ in 25 directions, 5 images with b=0 s/mm^2^, and resolution of 1×1×5 mm^3^ (7 minutes total). MT used 2D spoiled gradient echo ± MT pre-pulse (offset = 1.2 kHz), with 1×1×5mm^3^ voxels, for 8 minutes total. An additional T_1_-weighted image to calculate MTsat was also run. T_2_*-weighted acquisitions used a 2D Multi-Echo Recombined Gradient Echo (MERGE) with 3 echoes at 5, 10, and 15 ms, each with a resolution 0.6×0.6×4 mm^3^ without interpolation.

Total imaging time for all acquisitions together came to approximately 30 minutes after accounting for patient positioning, slice prescription, and high-order shimming. Advanced MRI scans underwent the same DICOM to BIDS conversion and manual inspection as the cMRI dataset described above. A summary of the varying scans and metrics in the advanced metric dataset are summarized in Table 1.

**Table 1.** Overview of MRI scan types, their descriptions, and derived features from these scans.

| **Metrics of Cord Compression** | **Description** | **Features** |
| --- | --- | --- |
| $T_2$-weighted Scan | $T_2$-w is a non-specific, common MRI scan type<sup>13</sup>. It was used for both community and advanced MRI datasets presented. | Spinal cord cross-sectional area, vertebral level segmentation |
| **Metrics of Axonal Integrity** |  |  |
| Diffusion Imaging** | Diffusion imaging captures evidence of directional diffusion of water in tissues<sup>14</sup>. They have shown promising results in the detection of CSM<sup>15</sup>. Fractional anisotropy has been found to be particularly sensitive<sup>15</sup>. | Fractional Anisotropy (FA) in spinal cord white matter |
| **Metrics of Demyelination** |  |  |
| T<sub>2</sub>*-weighted (T<sub>2</sub>*w) scan** | T<sub>2</sub>*w is a gradient signal echo image. T<sub>2</sub>*w images has been found to be useful in small lesion detection and the quantification of early degenerative disease <sup>16</sup> | Spinal cord white matter area and spinal cord grey matter area |
| Magnetization Transfer (MT) Scan** | MT scans are a useful scan for contrast augmentation <sup>17</sup>. MT scans have been found to detect myelin degradation in patients with CSM <sup>18</sup>. | Magnetization transfer ratio (MTR) in white matter |

\*\* used only in aMRI dataset

In addition to the imaging data, each patient had a clinical assessment where the modified Japanese Orthopedic Association (mJOA) score was recorded at the time of diagnosis. A patient’s mJOA score can range from 18 (no neurological deficits) to 0 (complete inability to move hands, complete loss of urinary sphincter control, and total loss of hand sensation)^19^. mJOA scores can be split into severity categories, designating mild (mJOA greater than or equal to 15), moderate (mJOA score between 14 and 12), or severe (mJOA of 11 or less) disease severity^20^. mJOA severity was used as the dependent variable in all machine learning models developed as part of this study. cMRI or aMRI dataset features were tested as independent variables for model predictions.

### Computational Tools Used

The versions of the tools used for these methods were as follows: scikit-learn v.0.23.2^21^, pandas v.1.1.3^22^, numpy v.1.19.2^23^, Spinal Cord Toolbox v.4.3^24^, matplotlib v.3.3.2^25^, and `Fsleyes v.0.34.2`^26^.

### MRI Scan Pre-processing

For both cMRI and aMRI records, raw DICOM data was extracted from each cMRI sequence and anonymized using the ‘`dcm2bids`’ package, converting them into a BIDS-compliant format^27^. Each patient record was manually inspected to confirm data integrity. Records were excluded if they contained motion artefacts, were mislabelled (i.e. the MRI were of the thoracic spine, rather than the cervical spine as labelled), and/or had an insufficient slice count (resulting in the inability for segmentation algorithms to make accurate estimates of spinal cord metrics).

### Spinal Cord MRI Analysis

Spinal Cord Toolbox (SCT), an all-in-one MRI processing package specializing in spinal cord MRI scans^28^, was used for image segmentation for both cMRI and aMRI datasets. Analysis metrics for both the community MRI and advanced MRI datasets were completed using the SCT’s `‘sct_deepseg_sc`’ command for the T2w, T2*w, and MT scan. SCT’s ‘`propseg`’ command was used for the DTI scan^24^. An example of these segmentations is shown in Figure 1.

**Figure 1.**
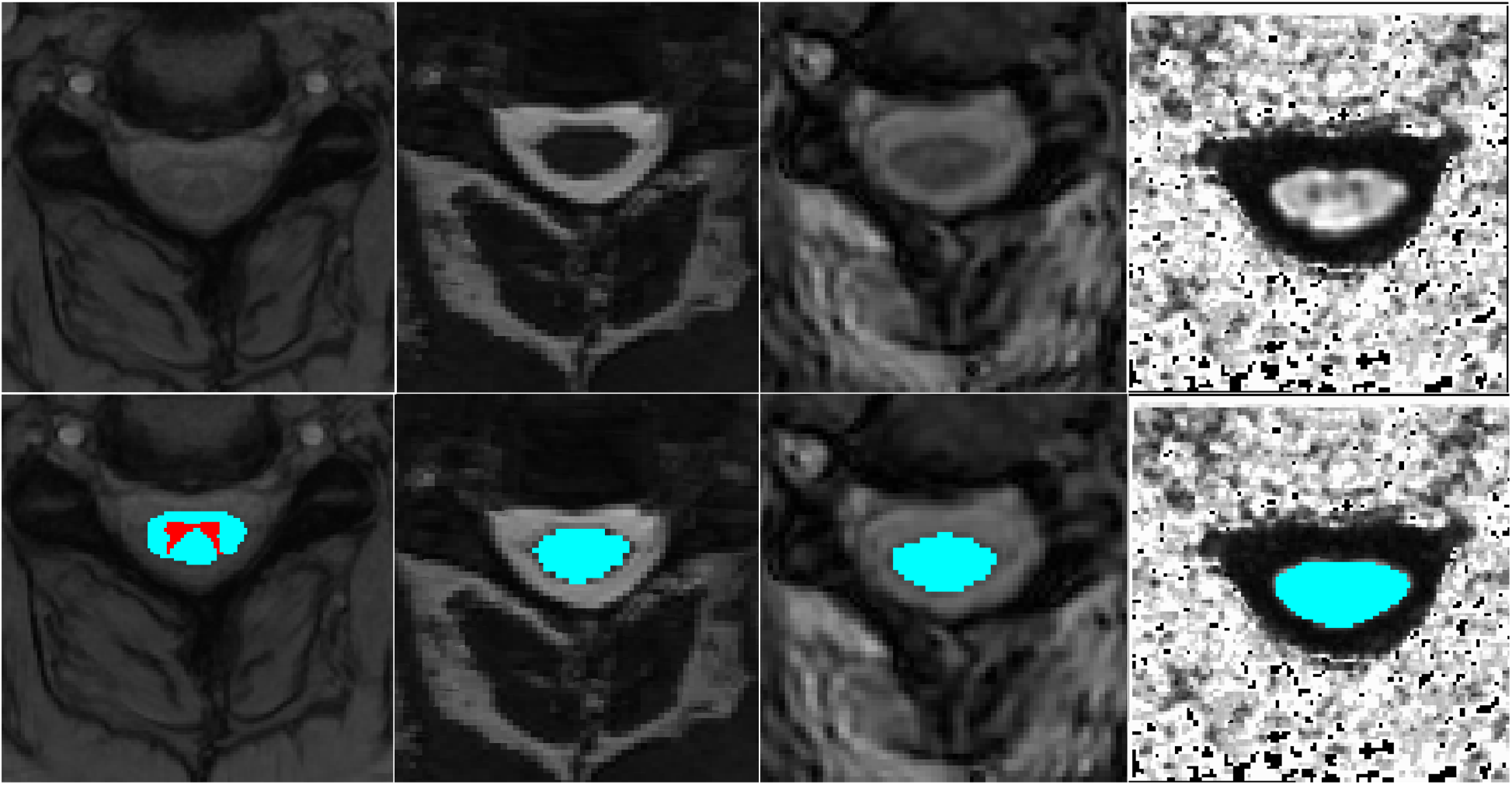
MRI scan type visualization and segmentation examples. Images were captured at the C3 level of a control patient. Top thumbnails show the image without segmentation, bottom images include segmentation. (Left to Right) T2*-weighted grey matter (red) and white matter (blue) scan, T2-weighted cross sectional area scan, Magnetization Transfer scan, Fractional Anisotropy in white matter.

MRI-derived metrics were then extracted from the T_2_-weighted and T_2_*-weighted segmentations using SCT’s ‘`sct_process_segmentation`’ function, creating the features used by our machine learning models. These features included the mean and standard deviation of clinically relevant metrics on a per-vertebrae level, including cross-sectional area, angle between centerline and axial slice in the anterior-posterior (AP) axis and right-left (RL) axis, distance between the major and minor axis of the cord (diameter) in the AP and RL axis, ratio of the focal distance over the major axis length (eccentricity), angle between AP axis of the spinal cord and AP axis of the image (orientation), spinal cord solidity, and spinal cord length^24^. This resulted in 9 features per vertebrae in the T_2_-weighted acquisition and 7 features per vertebrae in the T_2_*-weighted acquisition (as T_2_*-weighted does not contain Angle AP and Angle RL values). DTI and MT scans were also analyzed using SCT’s ‘`sct_extract_metric`’ script to generate the “maximum a posteriori (MAP)” for the Fractional Anisotropy in white matter and magnetization transfer ratio in white matter respectively ^24^.

For all MRI-derived metrics we calculated the overall mean, standard deviation, maximum value, minimum value, and the difference between maximum/minimum values (Δ) for all metrics across all vertebrae. The means for the cross sectional area of T_2_-weighted, spinal grey matter, spinal white matter, FA in white matter, and MTR were also included. Age and sex of the patient were also included as features in both datasets. This resulted in 53 features overall for the cMRI dataset and 149 features overall for an aMRI dataset.

### Management of missing clinical metrics/Excluded Metrics

Patients missing a recorded mJOA value or more than 20 percent of their imaging metrics were discarded. Remaining missing features were imputed using scikit-learn’s ‘`SimpleImputer’`. A specific breakdown of excluded patients can be found in Figure 2. Metrics from the C1 level in the T_2_-weighted and MT Scan, the C1-C3 + C7 level of the T_2_*-weighted scan, and the C1, C6, and C7 level of the DTI scan were excluded as SCT provided inconsistent and poor quality metric predictions for these vertebrae, this is likely a result of them appearing near the edge of the scan region.

**Figure 2.**
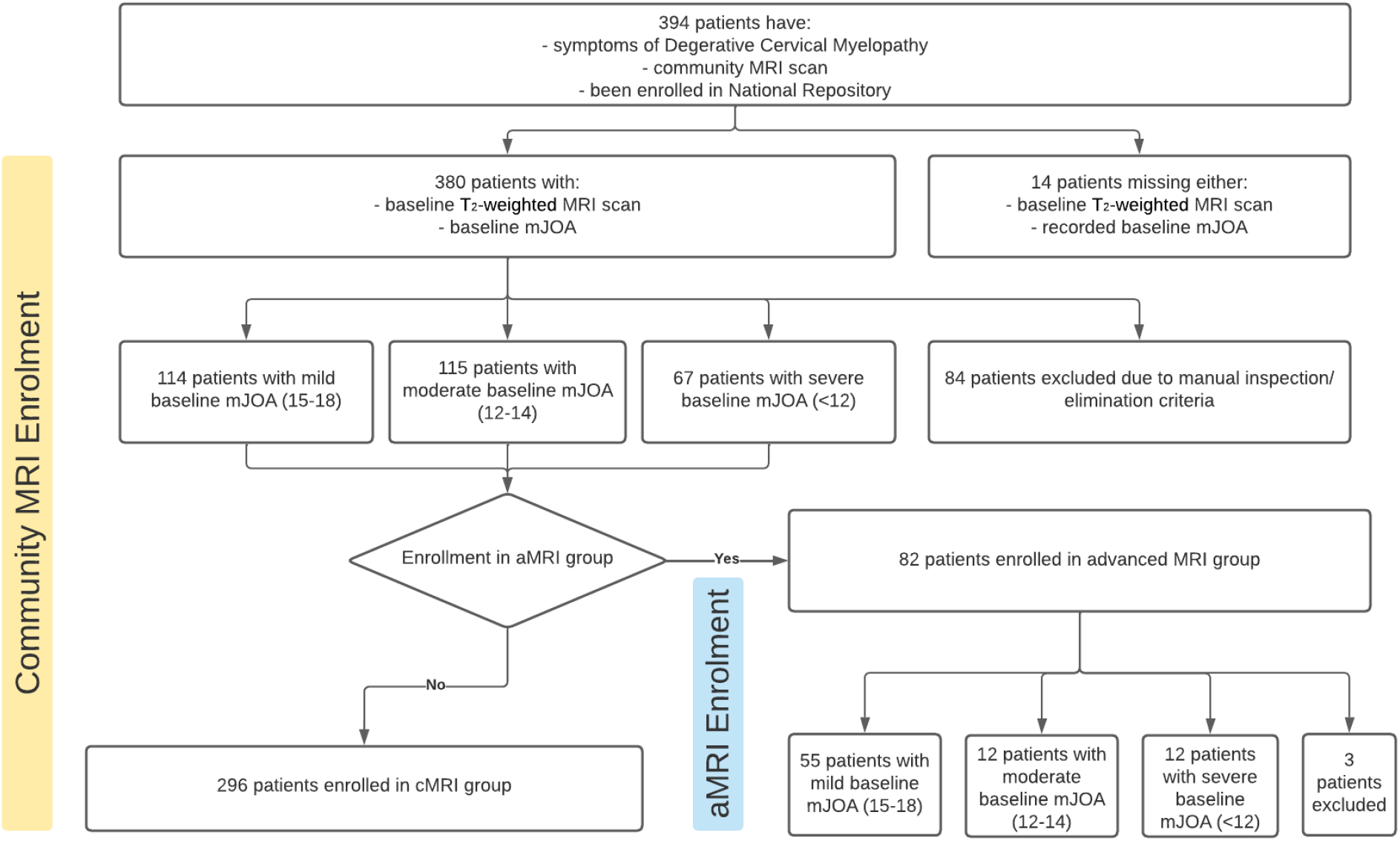
Breakdown of patient enrolment pathway and patient disease severity. Following excluded patients and scans, there were 296 cMRI patients. Of these patients, a subset of 82 patients were enrolled into the aMRI group. In the cMRI group, there were an equal number of mild and moderately severe patients (n=114 and n=115 respectively) with a decrease in the amount of severe mJOA patients (n=67). In the aMRI group, there was an abundance of mild severity patients (n=55) as compared to moderate and severe groups that were equal (n=12 per group).

### Prediction of baseline disease severity and Model Performance

Both cMRI and aMRI datasets were split into a training set consisting of 65 percent of the data and a testing set consisting of the remaining 35 percent. All features were scaled to a unit norm using scikit-learn’s ‘`StandardScaler`’ class. The mJOA score was also binned into 3 primary categories: mild severity (MJOA ≥ 15), moderate severity (MJOA = 12-14) and severe severity (MJOA < 12)^20^. This was done for two primary reasons: first, we cannot assume that every 1 point change in mJOA is equally significant, and secondly, these mJOA categories have been found to help clinicians assess disease severity ^20^.

Feature pre-processing methodologies were tested with and without Recursive Feature Elimination (RFE), with and without dimensionality reduction (using Principal Component Analysis (PCA) or Linear Discriminant Analysis (LDA) when dimensionality reduction was employed), with the resulting features being used to train and test Random Forest Classifier models^21^. The inclusion of RFE and PCA/LDA are important to ensure that the large feature set does not result in overfitting of the machine learning model and to better understand the importance of specific features. A pipeline of each permutation was run in the order they are listed, with each being fed into a Random Forest Classifier using default settings. Random Forest Classifiers were selected over Logistic Regression, Gradient Boosting Classifier, and Support Vector Classification models based on its performance in preliminary tests.

Using SciKit-Learn, accuracy (using ‘`accuracy_score`’), precision (using ‘`precision_score`’), and recall (using ‘‘`recall_score`’) scores were calculated for each model generated prior using the model’s predicted mJOA severity class and the patient’s true mJOA severity class^21^. A cross validation analysis was done for the ‘`accuracy_score`’ using SciKitLearn’s ‘`cross_val_score`’ script via stratified K fold cross-validation^21^. Both cMRI and aMRI datasets underwent the same analysis pipeline for accurate comparison.

## 3. Results

### Patient Enrollment

A total of 394 patients were enrolled from 2016 to 2021. Of the enrolled patients, 380 had a baseline mJOA recorded. Following manual inspection and excluded scans, there were 296 cMRI records included in this study.

A total of 82 patients were simultaneously enrolled in the collection of advanced, disease-specific metrics. 79 of these patients had a baseline mJOA and were included in our aMRI dataset.

A visualization of the enrolled patients can be seen in Figure 2.

### Predicting Baseline Severity in cMRI Scan Metrics

The cMRI dataset consisted of 53 features across 296 records The testing accuracy of the model training on data without RFE or LDA/PCA was 41.8 %, which decreased to 41.1% with RFE, and continued on a downward trend with the addition of LDA and PCA (Table 3). RFE identified 32 of the 53 features as the optimum number of features for the model’s performance (Appendix 1) The cMRI prediction model performs similarly in predicting mild and moderate disease severities (46.0% correct and 47.9% correct respectively) but misinterprets all severe mJOA patients as more mild (Figure 3).

**Table 2.**
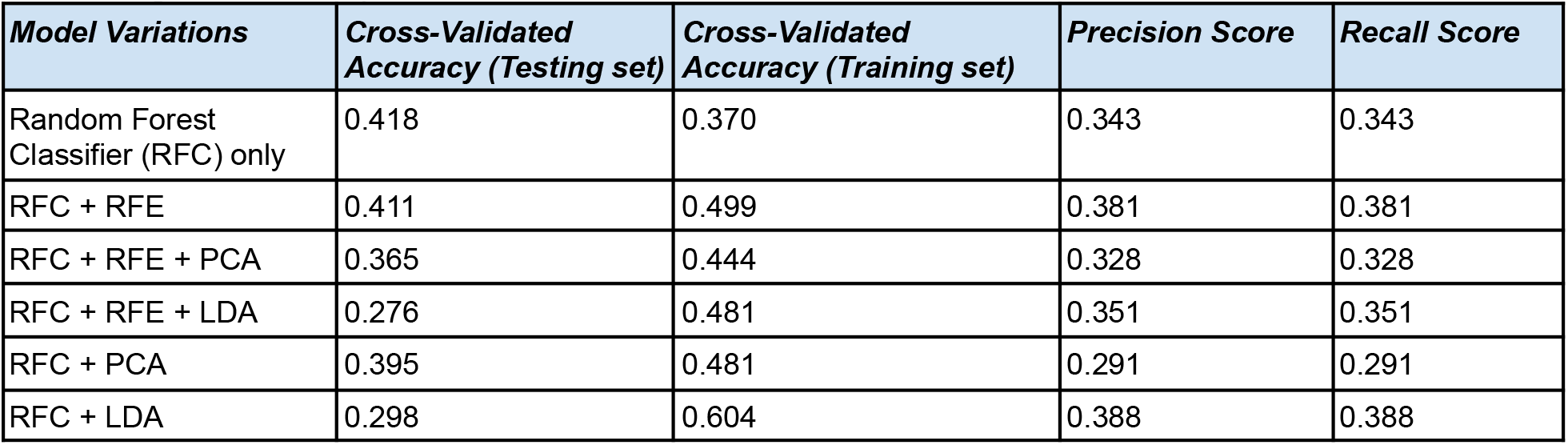
Summarized cross-validated accuracy, precision score, and recall score for tested model variations in the cMRI group. In the cMRI group, testing and training accuracies remained relatively the same with the exception of the RFC + LDA model which differed by 30.6% accuracy. The Random Forest Classifier model alone performed the best with an accuracy of 41.8%. The RFC + RFE + LDA model performed the worst with an accuracy of 27.6%. Precision and Recall scores followed the same trend and performed within 10% of their model’s respective accuracy.

**Table 3.**
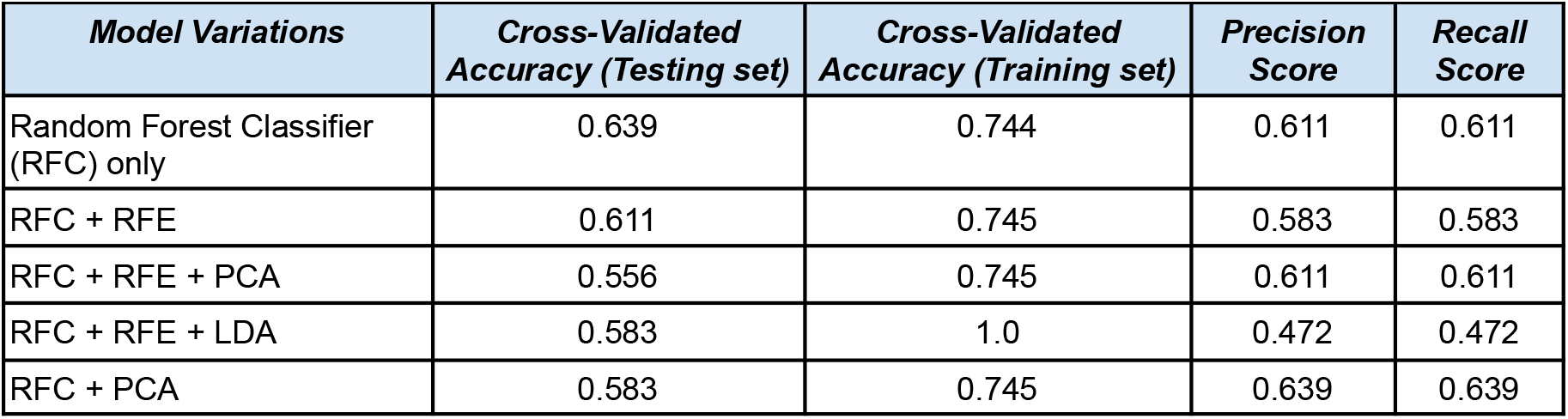
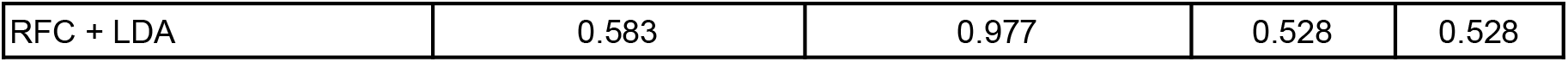
Summarized cross-validated accuracy, precision score, and recall score for tested model variations in the advanced metric dataset. In the aMRI group, testing and training accuracies remained similar with the exception of the RFC + RFE + LDA model and the RFC +LDA model which differed by 41.7% accuracy and 39.4% accuracy respectively. The Random Forest Classifier model alone performed the best with an accuracy of 63.9%. The RFC + RFE + PCA model performed the worst with an accuracy of 55.6 %. Precision and Recall scores followed the same trend and performed within 10% of their model’s respective accuracy.

| **Model Variations** | **Cross-Validated Accuracy (Testing set)** | **Cross-Validated Accuracy (Training set)** | **Precision Score** | **Recall Score** |
| --- | --- | --- | --- | --- |
| Random Forest Classifier (RFC) only | 0.639 | 0.744 | 0.611 | 0.611 |
| RFC + RFE | 0.611 | 0.745 | 0.583 | 0.583 |
| RFC + RFE + PCA | 0.556 | 0.745 | 0.611 | 0.611 |
| RFC + RFE + LDA | 0.583 | 1.0 | 0.472 | 0.472 |
| RFC + PCA | 0.583 | 0.745 | 0.639 | 0.639 |
| RFC + LDA | 0.583 | 0.977 | 0.528 | 0.528 |

**Figure 3.**
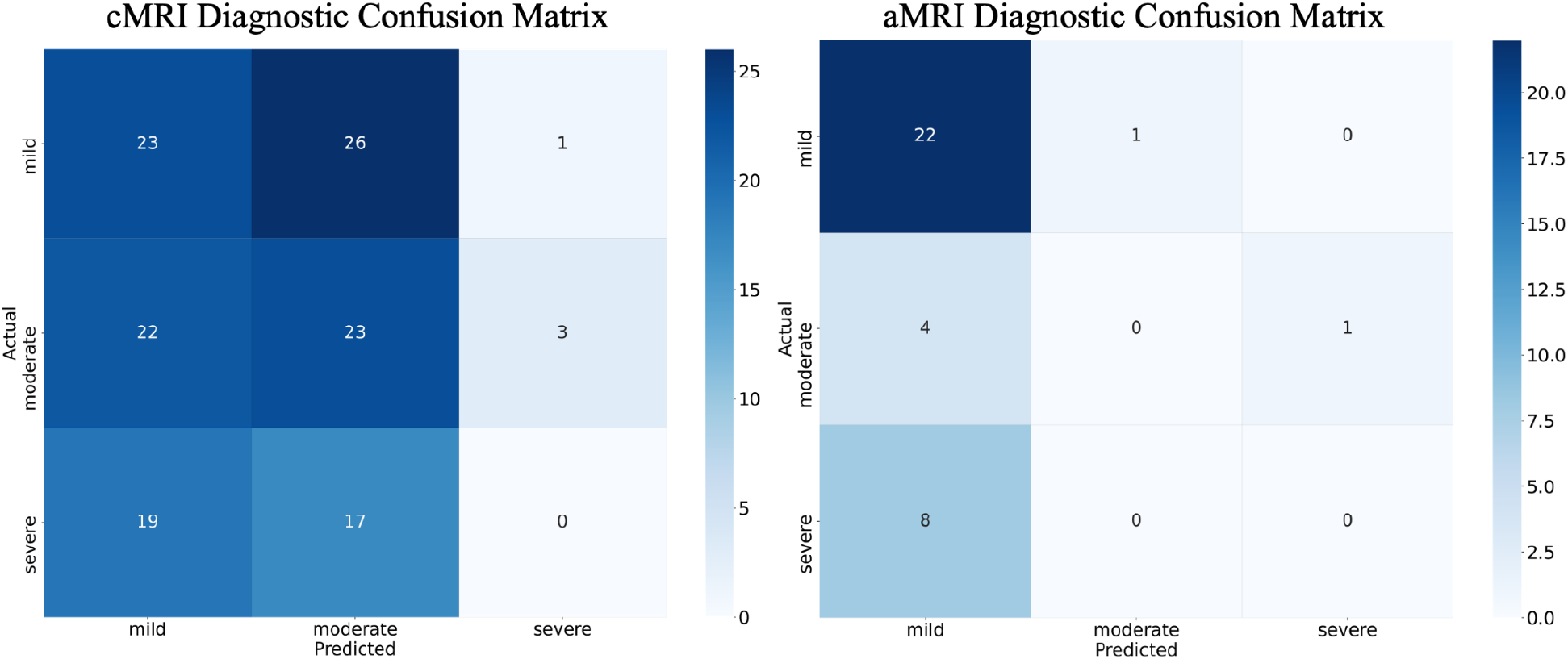
Confusion Matrix for cMRI and aMRI dataset model predictions across MJOA categories for baseline disease severity. The cMRI prediction model performed equally in predicting mild and moderate baseline disease severity with 23/50 correct and 23/48 correct respectively. The cMRI prediction model was not able to predict any of the 36 severe testing patients correctly. That being said, the model favours making moderate predictions (n=66) slightly more than mild predictions (n=64). It favours both mild and moderate predictions significantly more than severe predictions (n=4). The aMRI model predicts 22/23 mild patients correctly yet incorrectly the 5 moderate and 8 severe testing patients. It highly favours making mild predictions (n=34) as opposed to moderate or severe predictions (n=1 per group).

### Predicting Baseline Severity in aMRI Scan Metrics

The advanced MRI metrics consisted of 149 features accounting for metrics relating to the T_2_-weighted, T_2_*-weighted, DTI and MT scans. The initial testing accuracy of the model was 63.9% which decreased slightly with pre-processing permutation yet remained within ∼5.6% of the initial accuracy (Table 3). RFC+RFE+PCA was the one exception, dropping down to 55.6% accuracy. RFE identified 58 of the 149 features as the optimum number of features for the model’s performance (Appendix 2). The confusion matrix indicates that the model almost perfectly predicted mild disease severity (95.7% correct) yet predicted none of the moderate or severe patients correctly (Figure 3).

## 4.0 Discussion

In this work we compare the utility of community acquired diagnostic MRI scans with advanced MRI acquisitions to predict the severity of CSM at the time of diagnosis. The aMRI acquisition protocol was designed to capture evidence of traumatic spinal cord injury such as demyelination or axonal damage whereas community MRI acquisitions capture evidence of cord compression. The significance of this work lies with the novel implementation of machine learning models with aMRI acquisitions to better help physicians prioritize patients during triage and during diagnosis. This is important as MRI acquisition protocols for individuals with CSM have remained relatively unchanged since the 1980s while the available analysis tools and scans have improved^3^. Through testing the effectiveness of aMRI scans against cMRI scans, we are able to determine what role these scans can play to help physicians in the diagnosis of individuals with DCM. Furthermore, through analyzing which specific metrics had the most profound impact on disease severity prediction, we are better able to determine which scans should be prioritized by physicians.

We demonstrate that quantitative analysis of the aMRI dataset outperforms the predictive ability of community acquired MRI data in baseline disease severity prediction. Not only did the aMRI dataset have significantly improved accuracy (Figures 3, 4, and 5) of roughly 20% which was retained across pre-processing permutations, but also had consistently higher precision and recall scores in the diagnostic severity prediction. Furthermore, these improved accuracies were consistent across multiple tests and with a significantly smaller dataset (235 fewer scans) than the cMRI dataset. It is expected that with a larger aMRI dataset, this disparity between accuracy scores will only grow.

**Figure 4.**
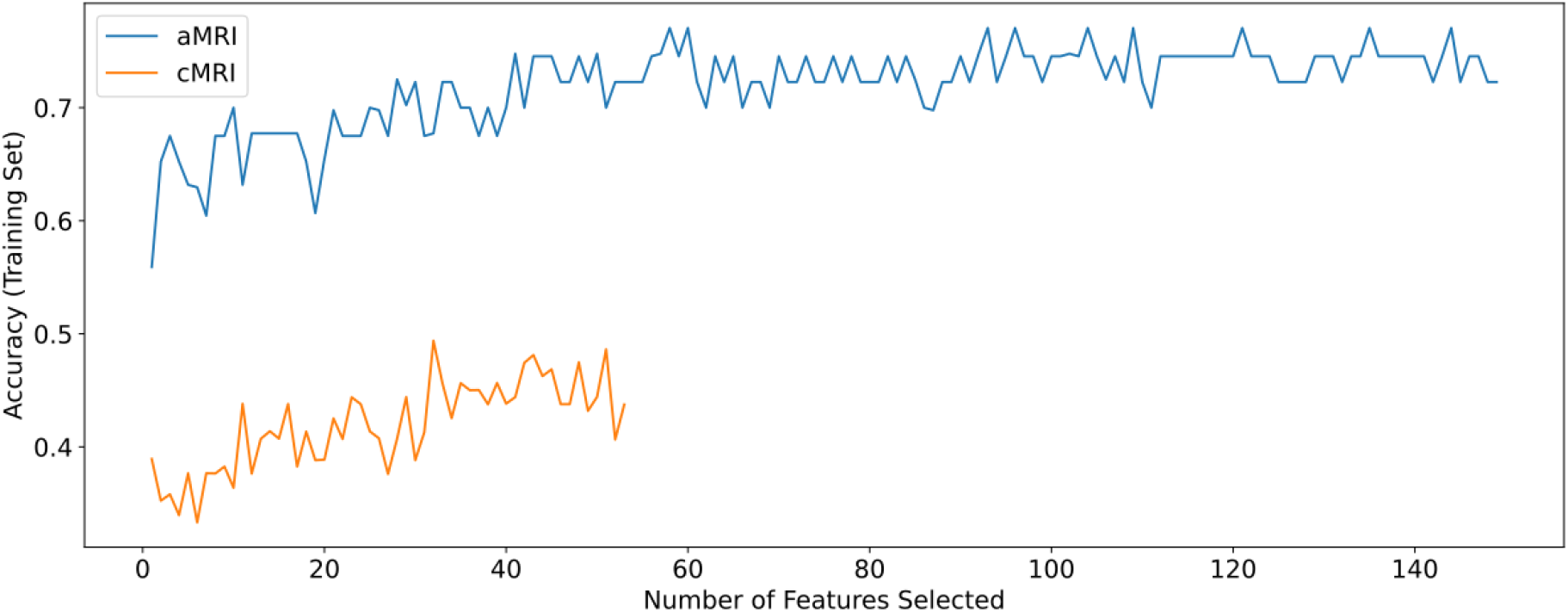
Model accuracy scores across the total feature set completed by recursive feature elimination in both the cMRI and aMRI groups. The cMRI group training accuracy began at ∼39% accuracy compared to the aMRI group ∼55% accuracy creating a 16% accuracy difference. Both models present an upward trend as more features are selected with the aMRI group beginning to plateau past 55 selected features. The cMRI group showed the greatest accuracy fluctuations around ∼20-37 selected features with the aMRI group having the greatest accuracy fluctuations around ∼0-25 selected features.

**Figure 5.**
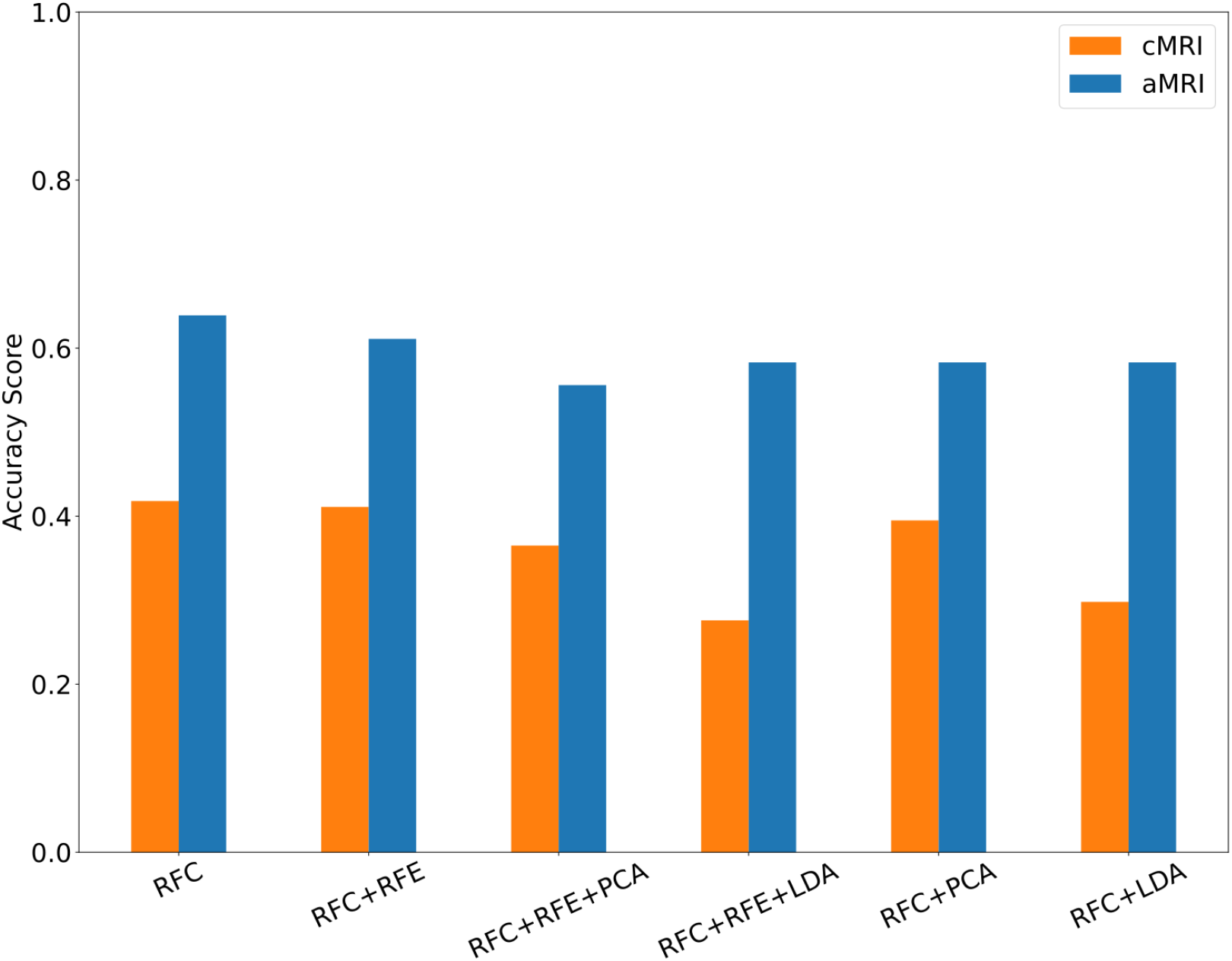
Accuracy score visualization for different model variations compared between the cMRI dataset and the aMRI dataset metrics. Both cMRI and aMRI dataset accuracies show minimal fluctuations between model variations. That being said, all the aMRI models outperform their cMRI counterparts. The largest difference between cMRI and aMRI accuracy scores is seen in the RFC+RFE+LDA model with a 30.7% prediction accuracy difference. The smallest difference between the cMRI and aMRI accuracy scores is seen in the RFC+PCA model with a 19.1% prediction accuracy difference.

Secondly, we demonstrate that cMRI acquisitions alone fail at accurately predicting disease severity when utilizing simple machine learning models. The cMRI dataset did not surpass 42% accuracy in any of the testing models despite the inclusion of RFE and PCA/LDA. Furthermore, the cMRI prediction model appears to act naively in predicting severe disease severity, failing to accurately identify any patients with severe DCM. We suspect that this is due to imbalance in the cMRI dataset, biasing predictions towards the more common mild and moderate DCM classes. It may also indicate that the cMRI-derived metrics are insufficient for simple machine learning models to function effectively, or that these simple models simply cannot work effectively with cMRI-derived metrics.

### Feature Utility Analysis

In the aMRI dataset, as seen in Figure 4, the model’s performance begins to plateau past 25 features and fully levels out with minor fluctuation beyond 55 features. This indicates that there are many possible combinations of features that can be used to predict disease severity, though many of these combinations are also redundant. Of the 58 features that were ranked as optimal by RFE, 20 out of 51 T2-weighted metrics were selected, 32 out of 77 T2*-weighted metrics were selected, 5 out of 9 DT scan metrics were selected, and 1 out of 19 MT scan metrics were selected. With more than half stemming from aMRI-derived feature, it is apparent that the accuracy benefits observed are at least in part due to the improved aMRI methodology. This is in line with prior research work that has shown grey matter volume loss may correlate with DCM severity^29^.

For example, both datasets took the T2w CSA area at lower vertebral levels (such as C6 and maximum values) which is in line with evidence to suggest that impactful compression is more often seen at lower cervical vertebral levels^30^. While both aMRI and cMRI datasets looked at the same features for the T2w scan, the aMRI dataset outlined 20 important features in comparison to the 38 features outlined by the cMRI dataset. This suggests that the new features introduced via the aMRI methodology were able to provide the information relied upon in the cMRI methodologies via another method, likely the result of the higher fidelity sequences obtained.

When comparing the confusion matrix between the cMRI and aMRI datasets (Figure 3), it is evident that the aMRI dataset outperformed the cMRI dataset in mild severity patients while the cMRI dataset outperformed the aMRI dataset in moderate severity patients. Neither of the two datasets performed well in predicting severe disease patients, likely due to both datasets being imbalanced against severe DCM severity.

### Limitations

A major limitation in this study was the lack of moderate and severe mJOA patient samples within the aMRI diagnostic dataset. Furthermore, this inequality in the number of each patient class within the aMRI set likely led the prediction model to act in a naive way, biased toward mild and moderate cases and neglecting severe ones. Another major limitation of this study was the variability in MRI scanning methodology for the community MRI dataset as compared to the aMRI set. The aMRI dataset had a strict acquisition protocol while the cMRI dataset was exposed to inter-clinic variability in acquisition methodology, likely leading to cMRI records having significantly more “noise” than equivalent aMRI records.

### Future Directions

Future focus will be on developing tools to predict long-term patient outcomes for both surgical and observational arms. To accomplish this, physician gathered metrics are currently being taken of patients to assess gait and balance, hand function, and global function of patients. These will be used as features in future models to determine if the gathering of these directed tests is useful in disease diagnosis or prognosis. Neural network models will be tested as well to assess whether they can bypass the limitations of the random forest models tested here. These models will also be able to accept MRI sequences directly, bypassing the need for extensive pre-processing and feature extraction. Finally, MRI sequence collection for both the cMRI and aMRI datasets is expected to continue, with future studies with these enlarged datasets aiming to artificially balance the dataset to avoid the imbalancing issues observed within our current results.

## Conclusion

The utilization of computer-based predictive modelling has a promising future in the prediction of baseline DCM disease severity. In this paper, the utilization of aMRI acquisitions such as T2*-weighted, DT imaging, and MT imaging have been shown to improve DCM severity prediction accuracy by 20 percent on average as compared to cMRI-derived metrics alone. Of the advanced acquisitions, T2*-weighted scans had the greatest impact in the increased accuracy score. Based on these results, it is apparent that the increased sequence fidelity obtained via the aMRI methodology appears to improve machine learning effectiveness, though improvements remain. These results were limited by the size and composition of our datasets, something which further data collection will alleviate. Such results are promising, but future work looking into neural network based models and improved patient metric selection must be completed before potential applications in a clinical setting can be considered.

## Data Availability

Due to patient privacy concerns, all data used and produced in the present study are only available upon request to the authors. Please contact Dr. David Cadotte for inquiries on this matter.

## Appendix 1: Community MRI Feature List in Diagnostic Severity Prediction

**Table 1.** The optimal and non-optimal list of features in the community MRI dataset following Recursive Feature Elimination

| Optimal Features |  |  | Non-Optimal Features |  |
| --- | --- | --- | --- | --- |
| T2w C4 CSA | T2w Diameter AP min | T2w Orientation $\Delta$ | Age | T2w Angle RL Std |
| T2w C5 CSA | T2w Diameter AP $\Delta$ | T2w Orientation mean | Sex | T2w Diameter AP max |
| T2w C6 CSA | T2w Diameter AP mean | T2w Solidity max | T2w C2 CSA | T2w Diameter RL min |
| T2w C7 CSA | T2w Diameter AP Std | T2w Solidity min | T2w C3 CSA | T2w Diameter RL Std |
| T2w CSA $\Delta$ | T2w Diameter RL max | T2w Solidity $\Delta$ | T2w Overall CSA mean | T2w Eccentricity min |
| T2w Angle AP max | T2w Diameter RL $\Delta$ | T2w Solidity mean | T2w CSA max | T2w Orientation min |
| T2w Angle AP min | T2w Diameter RL mean | T2w Length min | T2w CSA min | T2w Orientation Std |
| T2w Angle AP Std | T2w Eccentricity max | T2w Length mean | T2w CSA Std | T2w Solidity Std |
| T2w Angle RL max | T2w Eccentricity $\Delta$ | | T2w Angle AP $\Delta$ | T2w Length max |
| T2w Angle RL min | T2w Eccentricity mean |  | T2w Angle AP mean |  |
| T2w Angle RL $\Delta$ | T2w Eccentricity Std | | T2w Length Std | |
| T2w Angle RL mean | T2w Orientation max | | T2w Length $\Delta$ | |

\*The features are not placed in any particular order.

## Appendix 2: Advanced MRI Feature List in Diagnostic Severity Prediction

**Table 1.** The optimal and non-optimal list of features in the advanced MRI dataset following Recursive Feature Elimination

| Optimal Features |  | Non-Optimal Features |  |  |
| --- | --- | --- | --- | --- |
| T2w C6 CSA | GM Solidity mean | Age | T2w Length max | WM Diameter AP min |
| T2w CSA mean | GM Solidity Std | Sex | T2w Length min | WM Diameter AP $\Delta$ |
| T2w CSA max | GM Length min | MTR min tr_min | T2w Length $\Delta$ | WM Diameter AP mean |
| T2w CSA min | GM Length $\Delta$ | MTR Std tr_std | T2w Length mean | WM Diameter AP Std |
| T2w CSA Std | GM Length mean | T2w C2 CSA | GM C4 CSA | WM Diameter RL max |
| T2w Angle AP max | GM Length Std | T2w C3 CSA | GM C5 CSA | WM Eccentricity max |
| T2w Angle AP min | WM C4 CSA | T2w C4 CSA | GM C6 CSA | WM Eccentricity min |
| T2w Angle AP mean | WM CSA $\Delta$ | T2w C5 CSA | GM CSA $\Delta$ | WM Eccentricity $\Delta$ |
| T2w Angle AP Std | WM CSA Std | T2w C7 CSA | GM CSA max | WM Eccentricity mean |
| T2w Angle RL $\Delta$ | WM Diameter RL min | T2w CSA $\Delta$ | GM CSA Std | WM Eccentricity Std |
| T2w Diameter AP min | WM Diameter RL $\Delta$ | T2w Angle AP $\Delta$ | GM Diameter AP max | WM Orientation $\Delta$ |
| T2w Diameter AP mean | WM Diameter RL mean | T2w Angle RL max | GM Diameter AP mean | WM Solidity $\Delta$ |
| T2w Eccentricity min | WM Diameter RL Std | T2w Angle RL min | GM Diameter RL max | WM Solidity Std |
| T2w Eccentricity mean | WM Orientation max | T2w Angle RL mean | GM Diameter RL min | WM Length max |
| T2w Orientation max | WM Orientation min | T2w Angle RL Std | GM Diameter RL mean | WM Length min |
| T2w Orientation min | WM Orientation mean | T2w Diameter AP max | GM Eccentricity min | WM Length $\Delta$ |
| T2w Orientation $\Delta$ | WM Orientation Std | T2w Diameter AP $\Delta$ | GM Eccentricity $\Delta$ | WM Length mean |
| T2w Orientation mean | WM Solidity max | T2w Diameter AP Std | GM Eccentricity mean | WM Length Std |
| T2w Orientation Std | WM Solidity min | T2w Diameter RL max | GM Eccentricity Std | FA in WM max |
| T2w Length Std | WM Solidity mean | T2w Diameter RL min | GM Orientation max | FA in WM at C2 |
| GM CSA mean | WM:GM ratio mean | T2w Diameter RL $\Delta$ | GM Orientation min | FA in WM at C3 |
| GM CSA min | FA in WM mean | T2w Diameter RL mean | GM Orientation $\Delta$ | FA in WM at C4 |
| GM Diameter AP min | FA in WM min | T2w Diameter RL Std | GM Orientation mean | MTR mean |
| GM Diameter AP $\Delta$ | FA in WM Std | T2w Eccentricity max | GM Orientation Std | MTR at C2 mtr2 |
| GM Diameter AP Std | FA in WM $\Delta$ | T2w Eccentricity $\Delta$ | GM Length max | MTR at C3 |
| GM Diameter RL $\Delta$ | FA in WM at C5 | T2w Eccentricity Std | WM C5 CSA | MTR at C5 mtr5 |
| GM Diameter RL Std | MTR at C4mtr4 | T2w Solidity max | WM C6 CSA | MTR at C6 mtr6 |
| GM Eccentricity max | | T2w Solidity min | WM CSA mean | MTR $\Delta$ |
| GM Solidity max | | T2w Solidity $\Delta$ | WM CSA max | MTR max tr_max |
| GM Solidity min |  | T2w Solidity mean | WM CSA min |  |
| GM Solidity $\Delta$ | | T2w Solidity Std | WM Diameter AP max | |

\*The features are not placed in any particular order.

